# The prevalence of SARS-CoV-2 infection and long COVID in US adults during the BA.5 surge, June-July 2022

**DOI:** 10.1101/2022.09.04.22279588

**Authors:** Saba A Qasmieh, McKaylee M Robertson, Chloe A Teasdale, Sarah G Kulkarni, Heidi Jones, Margaret McNairy, Luisa N. Borrell, Denis Nash

## Abstract

Due to changes in SARS-CoV-2 testing practices, passive case-based surveillance may be an increasingly unreliable indicator for monitoring the burden of SARS-CoV-2, especially during surges.

We conducted a cross-sectional survey of a population-representative sample of 3,042 U.S. adults between June 30 and July 2, 2022, during the Omicron BA.5 surge. Respondents were asked about SARS-CoV-2 testing and outcomes, COVID-like symptoms, contact with cases, and experience with prolonged COVID-19 symptoms following prior infection. We estimated the weighted age and sex-standardized SARS-CoV-2 prevalence, during the 14-day period preceding the interview. We estimated age and gender adjusted prevalence ratios (aPR) for current SARS-CoV-2 infection using a log-binomial regression model.

An estimated 17.3% (95% CI 14.9, 19.8) of respondents had SARS-CoV-2 infection during the two-week study period–equating to 44 million cases as compared to 1.8 million per the CDC during the same time period. SARS-CoV-2 prevalence was higher among those 18-24 years old (aPR 2.2, 95% CI 1.8, 2.7) and among non-Hispanic Black (aPR 1.7, 95% CI 1.4, 2.2) and Hispanic (aPR 2.4, 95% CI 2.0, 2.9). SARS-CoV-2 prevalence was also higher among those with lower income (aPR 1.9, 95% CI 1.5, 2.3), lower education (aPR 3.7 95% CI 3.0,4.7), and those with comorbidities (aPR 1.6, 95% CI 1.4, 2.0). An estimated 21.5% (95% CI 18.2, 24.7) of respondents with a SARS-CoV-2 infection more than 4 weeks prior reported long COVID symptoms.

The inequitable distribution of SARS-CoV-2 prevalence during the BA.5 surge will likely drive inequities in the future burden of long COVID.

## INTRODUCTION

The COVID-19 pandemic continues to cause significant morbidity and mortality both in the United States (U.S.) and globally. While vaccines and boosters against SARS-CoV-2 have shown dramatic effectiveness in reducing COVID-19 hospitalizations and deaths^1^, the circulation of more transmissible SARS-CoV-2 sub-variants and waning immunity underscores the importance of continued monitoring of SARS-CoV-2 burden. At the writing of this report (August 2022), BA.5 is the predominant subvariant circulating in the U.S^2^. Given the dynamic and uncertain nature of the virus at this time it is imperative that useful, robust, and proactive SARS-CoV-2 monitoring systems be maintained to characterize both short- and long-term sequelae of the pandemic (i.e., long COVID).

While the virus and its impact have evolved substantially, the current approaches to COVID-19 public health surveillance in the U.S. has not evolved to keep pace, making tracking of the true burden of SARS-CoV-2 increasingly challenging. Declines in SARS-CoV-2 screening and diagnostic testing combined with the increased use of at-home rapid antigen tests^3^ (which are not generally captured via routine case-based surveillance), are likely resulting in an increasing underestimation of the true case burden of infection^4^. Low testing and testing driven mostly by symptoms and exposure may also inflate test positivity rates reported by testing sites relative to those in the general population^5^. In addition, the extent of incomplete reporting of positive tests at local, state, and national level is yet to be evaluated. Case data from passive case-based surveillance reflect a non-representative sample of individuals who present for testing^6^ which further complicates the interpretation of trends in case numbers. Population-based surveys can, therefore, complement both routinely collected case-based data to inform public health mitigation measures^7^.

The high exposure to transmissible sub-variants is also likely to contribute to a growing number of individuals currently experiencing long COVID. Long COVID, or the recurring or ongoing symptoms or clinical findings four or more weeks after SARS-CoV-2 infection is likely affecting millions of Americans. Recent estimates from the Centers for Disease Control and Prevention (CDC) show that at least 1 in 5 adults with prior SARS-CoV-2 infection experience long COVID^8^. There are currently only a few published studies^9^ from the U.S. measuring current infections combining individual-level information on demographics, prior infection history, vaccination status, long COVID, education status, and income. Using a population-representative survey, this study aimed to estimate the prevalence of SARS-CoV-2, characterize factors associated with testing and infection, and the prevalence of long COVID among U.S. adults with a prior SARS-CoV-2 infection.

## METHODS

### Survey-based estimation of SARS-CoV-2 prevalence

We conducted a cross-sectional survey, in English and Spanish, between June 30 and July 2, 2022, of 3,042 adult U.S. residents via landlines (IVR) and mobile phones (SMS text). Potential participants were randomly selected from a sampling frame of U.S adults. To create weights representative of the US-population 18 years or older, we used an iterative weighting method, raking, to marginal proportions of race, ethnicity, age, self-identified sex, and education by US region based on the 2020 US census^10^. Further details on the survey design, sampling and weighting are provided in Appendix 1.

The survey questionnaire (Appendix 2) ascertained SARS-CoV-2 testing results of viral PCR, rapid antigen and/or at-home rapid diagnostic tests taken in the 14 days prior to the survey (June 16 - July 2). During the same time period, the BA.5 Omicron subvariant rose from an estimated 29.1% of reported cases to 57%^2^. The survey captured information on COVID-19 symptoms, as well as known close contacts with a confirmed or probable case of SARS-CoV-2 infection. COVID-19 symptoms included any of the following: fever of >100°F, cough, runny nose and/or nasal congestion, shortness of breath, sore throat, fatigue, muscle/body aches, headaches, loss of smell/taste, nausea, vomiting and/or diarrhea^11^. Participants were also asked about vaccination status (but not type(s) or date(s) of vaccination), comorbidities that increase vulnerability to severe COVID-19, and prior history of SARS-CoV-2 infection/COVID.

#### Point Prevalence estimation

Information gathered from respondents was used to estimate the number and proportion of respondents who likely had SARS-CoV-2 infection during the study period based on the following mutually exclusive, hierarchical case classification: 1) Confirmed case: self-report of one or more positive tests with a health care or testing provider; 2) Probable case: self-report of a positive test result exclusively on at-home rapid tests (i.e. those that were not followed up with confirmatory diagnostic testing with a provider); or 3) Possible case: self-report of COVID-like symptoms AND a known epidemiologic link (close contact) to one or more laboratory confirmed or probable (symptomatic) SARS-CoV-2 case(s)^11^ in a respondent who reported never testing or only testing negative during the study period.

#### The intersection of vaccine- and infection-induced immunity

We combined information on current vaccination status along with that on prior COVID infections (as of June 15, 2022) to classify those who were fully vaccinated and those who were also boosted at least once (fully vaccinated/boosted) with a history of prior COVID as having, ‘hybrid immunity’ against severe COVID; those who were fully vaccinated or boosted at least once with no history of prior COVID were classified as having ‘vaccine-induced immunity only’; those who were not fully vaccinated but had a history or prior COVID were classified as having ‘infection-induced immunity only’; and those who were neither vaccinated/boosted nor had a history of COVID were classified as having ‘no prior immunity’ (SARS-CoV-2 naive).

#### Long COVID definition

We used a question routinely used by the Office of National Statistics in the United Kingdom to define and assess the burden of long COVID^7^. Respondents in our survey who reported a history of prior COVID were asked “Would you describe yourself as having ‘long COVID’, that is you experienced symptoms such as fatigue, difficulty concentrating, shortness of breath more than 4 weeks after you first had COVID-19 that are not explained by something else?”. The point prevalence of long COVID was estimated among those with prior SARS-CoV-2 infection as the proportion responding affirmatively. Respondents whose most recent SARS-CoV-2 infection was within the past 4 weeks were classified as not having long COVID, to avoid conflation of symptoms of acute illness, and to align with the definition of long COVID, which was assessed more than four weeks after infections.

#### SARS-CoV-2 routine testing and case surveillance data

We used publicly available, daily aggregated data on the number of SARS-CoV-2 diagnostic tests and positive results through July 2, 2022^12^, to compare the number of tests and number positives reported to the CDC during the study period.

#### Statistical Analysis

We estimated the prevalence of SARS-CoV-2 and long COVID by socio- demographic characteristics, region, vaccination status, comorbidity, and prior COVID-19 status. Survey weights were applied to generate population-representative numbers and estimates of the proportion who were SARS-CoV-2 positive at any time during the study period along with 95% confidence interval (95% CI). We applied these weighted sample proportions and 95% CIs to the 254,297,978 US residents ≥18 years to obtain estimates of the absolute number of adults with SARS-CoV-2 infection and long COVID. Pearson’s chi-squared test was performed to assess associations between respondent characteristics and testing status. We used direct standardization to calculate age and sex adjusted prevalence estimates using the U.S. 2020 Census. Crude and age- and gender-adjusted prevalence ratios and 95% CIs were calculated using a log-binomial model. We used sex for standardization and gender for adjustment since our survey captured only gender. For standardization, we had to assume that reported gender was reported sex.

The study protocol was approved by the Institutional Review Board at the City University of New York (CUNY IRB 2022-0407).

## RESULTS

### Prevalence

The weighted characteristics of survey participants are shown in Table 1. We estimate that 17.3% (95% CI 14.9, 19.8) of the approximately 254,297,978 million U.S. adults had SARS-CoV-2 between 16-June and 2-July 2022, which corresponds to about 43,993,550 adults (95% CI 37,890,399 - 50,351,000). The SARS-CoV-2 prevalence estimate of 17.3% includes: 1) 10.0% (95% CI 7.6, 12.3) of respondents who tested positive based on one or more tests with a healthcare or testing provider, 2) 4.8% (95% CI 3.8,5.7) who were positive exclusively based on one or more at-home rapid tests; and 3) 2.6% (95% CI 1.8, 3.4) who met the definition for possible SARS-CoV-2 infection. The test positivity rate among those who tested with a healthcare or testing provider was 33.9% (95% CI 27.6, 40.1). We estimated that between 16-June and 2-July 2022, 9,435,413 tests were reportable to CDC, of which 1,805,033 were positive (i.e., 19.1% positivity rate among testers).

**Table 1.** Characteristics for survey respondents point prevalence of SARS-CoV-2, US adults, July 2022.

|  | Total | SARS-CoV-2 cases | Crude prevalence of SARS-CoV-2 infection† | Age and sex direct-standardized prevalence of SARS-CoV-2 infection** | Crude prevalence ratio (PR) | Adjusted prevalence ratio (aPR)*** |
| --- | --- | --- | --- | --- | --- | --- |
|  | N (%) | No. (%) | % (95% CI) | % (95% CI) | PR (95% CI) | aPR (95% CI) |
| **Total** | 3042 (100%) | 527 (100%) | 17.3 (14.9, 19.8) |  |  |  |
| **Testing** |  |  |  |  |  |  |
| Confirmed |  | 303 (10.0) | 10.0 (7.6, 12.3) |  |  |  |
| Probable |  | 145 (4.8) | 4.8 (3.8, 5.7) |  |  |  |
| Possible cases (non-testers/negatives) |  | 79 (2.6) | 2.6 (1.8, 3.4) |  |  |  |
| **Age** |  |  |  |  |  |  |
| 18-24 | 365 (12.0) | 161 (30.5) | 44.1 (32.5, 55.6) | 41.3 (32.7, 50.4) | 2.3 (1.9, 2.8) | 2.2 (1.8, 2.7) |
| 25-34 | 547 (18.0) | 105 (19.8) | 19.1 (13.2, 25.1) | 20.3 (14.4, 27.7) | Ref | Ref |
| 35-44 | 495 (16.3) | 79 (14.9) | 15.9 (10.5, 21.2) | 15.6 (10.8, 21.9) | 0.8 (0.6, 1.1) | 0.8 (0.6, 1.0) |
| 45-54 | 498 (16.4) | 63 (11.9) | 12.6 (8.0, 17.2) | 12.4 (8.4, 18.0) | 0.7 (0.5, 0.9) | 0.6 (0.5, 0.8) |
| 55-64 | 508 (16.7) | 52 (9.8) | 10.2 (7.4, 12.9) | 10.3 (7.8, 13.4) | 0.5 (0.4, 0.7) | 0.5 (0.4, 0.7) |
| 65+ | 629 (20.7) | 69 (13.1) | 11.0 (8.9, 13.0) | 10.7 (8.8, 12.9) | 0.6 (0.4, 0.8) | 0.5 (0.4, 0.7) |
| **Gender** |  |  |  |  |  |  |
| Male | 1443 (47.5) | 321 (60.9) | 22.2 (17.9, 26.6) | 22.9 (19.3, 26.9) | 1.8 (1.5, 2.1) | 1.9 (1.6, 2.2) |
| Female | 1516 (50.0) | 185 (35.1) | 12.2 (9.9, 14.5) | 12.2 (10.1, 14.7) | Ref | Ref |
| Non-binary | 82 (2.7) | 21 (4.0) | 25.5 (12.1, 38.9) | 22.7 (13.8, 35.1) | 2.1 (1.4, 3.1) | 1.5 (1.0, 2.2) |
| **Race/Ethnicity** |  |  |  |  |  |  |
| Black NH | 350 (11.5) | 86 (16.3) | 24.5 (17.6, 31.4) | 22.7 (17.4, 29.2) | 2.1 (1.7, 2.7) | 1.7 (1.4, 2.2) |
| White NH | 1794 (59.0) | 206 (39.1) | 11.5 (9.9, 13.1) | 11.9 (10.0, 14.0) | Ref | Ref |
| Hispanic | 460 (15.1) | 172 (32.6) | 37.4 (27.0, 47.7) | 25.8 (19.5, 33.3) | 3.3 (2.7, 3.9) | 2.4 (2.0, 2.9) |
| Asian/Pacific Islander | 171 (5.6) | 34 (6.4) | 19.7 (5.5, 33.9) | 16.3 (8.3, 29.5) | 1.7 (1.2, 2.4) | 1.1 (0.8, 1.5) |
| Other | 268 (8.8) | 30 (5.6) | 11.1 (5.2, 16.9) | 9.7 (5.3, 17.2) | 1.0 (0.7, 1.4) | 0.8 (0.6, 1.2) |
| **Years of education** |  |  |  |  |  |  |
| Some HS and below | 353 (11.6) | 194 (36.9) | 55.1 (43.4, 66.8) | 41.1 (30.4, 52.6) | 4.9 (4.0, 6.1) | 3.7 (3.0, 4.7) |
| HS Grad | 830 (27.3) | 94 (17.4) | 11.3 (8.2, 14.3) | 11.7 (8.7, 15.4) | Ref | Ref |
| Some college | 934 (30.7) | 112 (21.2) | 12.0 (9.3, 14.7) | 12.2 (9.5, 15.5) | 1.1 (0.8, 1.4) | 1.0 (0.8, 1.4) |
| College grad and above | 925 (30.4) | 127 (24.2) | 13.8 (11.4, 16.2) | 14.5 (11.9, 17.5) | 1.2 (1.0, 1.6) | 1.2 (1.0, 1.6) |
| **Household income** |  |  |  |  |  |  |
| Below 20K | 561 (18.4) | 187 (35.6) | 33.4 (24.6, 42.2) | 28.2 (22.9, 34.1) | 2.3 (1.9, 3.0) | 1.9 (1.5, 2.3) |
| 20,000 - 60,000 | 941 (31.0) | 124 (23.6) | 13.2 (9.9, 16.5) | 12.7 (9.9, 16.1) | 0.9 (0.7, 1.2) | 0.9 (0.7, 1.2) |
| 60,000 - 100,000 | 568 (18.7) | 81 (15.4) | 14.2 (10.6, 17.8) | 14.5 (11.1, 18.8) | Ref | Ref |
| Above 100,000 | 460 (15.1) | 69 (13.0) | 14.9 (10.4, 19.5) | 14.4 (10.4, 19.6) | 1.0 (0.8, 1.4) | 1.0 (0.7, 1.3) |
| Prefer not to answer | 443 (15.6) | 51 (9.7) | 11.5 (7.0, 16.0) | 15.3 (9.5, 23.9) | 0.8 (0.6, 1.1) | 0.8 (0.6, 1.1) |
| DK | 69 (2.3) | 15 (2.8) | 21.7 (6.1, 37.3) | 27.6 (15.4, 44.4) | 1.5 (0.9, 2.5) | 1.4 (0.8, 2.2) |
| **Employed** |  |  |  |  |  |  |
| Yes | 1472 (48.4) | 335 (63.5) | 22.7 (18.6, 26.8) | 19.9 (17.2, 22.9) | Ref | Ref |
| No/DK | 1570 (51.6) | 193 (36.6) | 12.3 (9.6, 14.9) | 12.9 (9.4, 17.3) | 0.5 (0.5, 0.6) | 0.6 (0.5, 0.7) |
| **Geographic region** |  |  |  |  |  |  |
| Northeast | 541 (17.8) | 112 (21.3) | 20.7 (15.1, 26.3) | 19.7 (15.8, 24.3) | Ref | Ref |
| South | 1153 (37.9) | 181 (34.3) | 15.7 (12.2, 19.2) | 15.8 (12.7, 19.4) | 0.8 (0.6, 0.9) | 0.8 (0.6, 0.9) |
| Midwest | 630 (20.7) | 75 (14.2) | 11.9 (7.6, 16.1) | 11.8 (7.8, 17.4) | 0.6 (0.4, 0.8) | 0.6 (0.5, 0.8) |
| West | 718 (23.6) | 160 (30.3) | 22.2 (15.7, 28.7) | 21.9 (17.4, 27.0) | 1.1 (0.9, 1.4) | 1.1 (0.9, 1.4) |
| **Vaccination status** |  |  |  |  |  |  |
| Boosted | 1610 (52.9) | 374 (70.9) | 23.2 (19.4, 27.0) | 24.1 (20.9, 27.6) | 3.5 (2.4, 4.7) | 3.0 (2.1, 4.2) |
| Fully vaccinated not boosted | 476 (15.6) | 33 (6.2) | 6.9 (4.0, 9.8) | 6.4 (4.2, 9.7) | Ref | Ref |
| Not vaccinated | 957 (31.5) | 121 (22.9) | 12.6 (8.6, 16.9) | 11.9 (8.7, 15.9) | 1.8 (1.3, 2.6) | 1.5 (1.1, 2.2) |
| **Prior COVID since March 2020** |  |  |  |  |  |  |
| Never | 1554 (51.1) | 116 (21.9) | 7.4 (5.7, 9.2) | 7.6 (5.8, 10.0) | 0.2 (0.2, 0.3) | 0.2 (0.2, 0.3) |
| Once | 808 (26.6) | 272 (51.6) | 33.6 (26.9, 40.2) | 26.7 (22.6, 31.3) | Ref | Ref |
| More than once | 343 (11.3) | 89 (16.9) | 25.9 (19.7, 31.1) | 28.0 (22.5, 34.3) | 0.8 (0.6, 0.9) | 0.8 (0.7, 1.0) |
| Never tested positive but think they had COVID | 335 (11.0) | 51 (9.6) | 15.1 (9.5, 20.7) | 16.1 (10.9, 23.1) | 0.5 (0.3, 0.6) | 0.5 (0.4, 0.7) |
| **Hybrid immunity** |  |  |  |  |  |  |
| Vaccine- and infection-induced | 987 (32.4) | 315 (59.7) | 31.9 (26.6, 37.2) | 29.2 (25.3, 33.3) | Ref | Ref |
| Vaccine-induced only | 1099 (36.1) | 92 (17.4) | 8.3 (6.2, 10.5) | 8.3 (6.0, 11.6) | 0.3 (0.2, 0.3) | 0.3 (0.3, 0.4) |
| Infection-induced only | 501 (16.5) | 97 (18.3) | 19.3 (12.5, 26.0) | 17.0 (12.4, 22.8) | 0.6 (0.5, 0.7) | 0.6 (0.5, 0.8) |
| No immunity | 456 (15.0) | 24 (4.6) | 5.3 (2.3, 8.2) | 5.8 (3.2, 10.2) | 0.2 (0.1, 0.2) | 0.2 (0.1, 0.3) |
| **Comorbidities** |  |  |  |  |  |  |
| Yes | 1104 (36.3) | 316 (59.9) | 28.6 (23.5, 33.6) | 28.8 (24.8, 33.2) | 2.6 (2.2, 3.1) | 2.8 (2.4, 3.3) |
| No | 1938 (63.7) | 212 (40.2) | 10.9 (8.7, 13.1) | 11.4 (9.3, 13.9) | Ref | Ref |
| **Any vulnerability\*** |  |  |  |  |  |  |
| Yes | 2106 (69.2) | 401 (76.1) | 19.0 (15.9, 22.2) | 19.6 (16.9, 22.5) | 1.4 (1.2, 1.7) | 1.6 (1.4, 2.0) |
| No | 936 (30.8) | 126 (23.9) | 13.5 (10.1, 16.9) | 12.6 (9.9, 16.1) | Ref | Ref |
| **Health insurance** |  |  |  |  |  |  |
| Yes | 2398 (78.8) | 438 (83.0) | 18.3 (15.4, 21.1) | 19.1 (16.7, 21.7) | Ref | Ref |
| No | 644 (21.2) | 89 (17.0) | 13.9 (9.5, 18.3) | 12.9 (9.2, 17.9) | 0.8 (0.6, 0.9) | 0.6 (0.5, 0.8) |

\*Aged 65 or older OR >1 comorbidity OR unvaccinated
\*\*Direct standardized for the age and sex groupings based in the 2020 U.S. census, except for age (standardized for sex only) and gender (standardized for age only)
\*\*Models adjusted for gender and age

Sex-standardized SARS-CoV-2 prevalence was higher among 18–24-year-olds (41.3%, 95% CI 32.7, 50.4), while age-standardized prevalence was higher among males (22.9%, 95% CI 19.3, 26.9). Age- and sex-standardized prevalence was higher among Hispanic adults (25.8%, 95% CI 19.5, 33.3), and adults with HS education level or below (41.1%, 95% CI 30.4, 52.6). Age- and sex-standardized prevalence was also higher among adults in the lowest category of annual household income below $20,000 (28.2%, 95% CI 22.9, 34.1). Regional differences in prevalence were also observed, with higher standardized prevalence reported in the West region of the U.S. (21.9%, 95% CI 17.4, 27.0), followed by the Northeast region (19.7%, 95% CI 15.8, 24.3).

### Hybrid immunity

Among the 32.4% (95% CI 30.0, 34.9) of those who were either vaccinated/boosted and who also had SARS-CoV-2 infection in the past (hybrid immunity), the age- and sex-standardized prevalence was 29.2% (95% CI 25.2, 33.3). SARS-CoV-2 prevalence was 8.3% (95% CI 6.0, 11.6) among respondents with vaccine-induced immunity but not immunity from a prior SARS-CoV-2 infection in the past, 17.0% (95% CI 12.4, 22.8) among those who have infection-induced immunity but were never vaccinated, and 5.8% (95% CI 3.2, 10.2) among respondents who were neither vaccinated/boosted nor had prior SARS-CoV-2 infection.

### Vulnerability to severe COVID-19

The estimated prevalence of SARS-CoV-2 was 11.9% (95% CI 8.7, 15.9) among unvaccinated, 10.7% (95% CI 8.8, 12.9) among those 65 years or older, and 28.8% (95% CI 24.8, 33.2) among respondents who reported a comorbidity. Among those with any of these vulnerabilities to severe COVID-19 (age ≥65, comorbidities, unvaccinated), 19.6% (95% CI 16.9, 22.5) had SARS-CoV-2 infection.

### Testing

Just under half (42.2%) of the adults in our sample reported receiving any SARS-CoV-2 test during the study period. About 2.2% of the sample reported testing with a health or testing provider only, 12.3% tested exclusively using an at-home rapid antigen test, and 27.3% tested both with a provider and at home. Differences were observed between testers and non-testers (Table 2) with testers more likely to be between 18 and 24 years old, and less likely to be above 55 years old. Testers were more likely to be Hispanic (22.1%), have less than a HS degree (19.7%), be employed (52.5%) and live in the Northeast region (21.0%). Testers were also more likely to have received a booster (62.3%) and have hybrid immunity (45.6%).

**Table 2.** Characteristics of survey respondents by testing status, US adults, July 2022.

|  | Total | Non-Testers | Testers (any) | Test with provider only | Test at home only | Test with both provider and at home |  |
| --- | --- | --- | --- | --- | --- | --- | --- |
|  | No. (%) | No. (%) | No. (%) | No. (%) | No. (%) | No. (%) | p-value** |
| **Total** | 3042 (100) | 1758 (57.8) | 1284 (42.2) | 66 (2.2) | 389 (12.3) | 829 (27.3) |  |
| **Age** |  |  |  |  |  |  |  |
| 18-24 | 365 (12.0) | 132 (7.5) | 233 (18.1) | 15 (22.6) | 27 (6.9) | 191 (23.1) | <0.0001 |
| 25-34 | 547 (18.0) | 292 (16.6) | 255 (19.8) | 5 (7.7) | 72 (18.5) | 178 (21.5) |  |
| 35-44 | 495 (16.3) | 329 (18.7) | 166 (12.9) | 14 (21.0) | 46 (11.7) | 107 (21.5) |  |
| 45-54 | 498 (16.4) | 276 (15.7) | 222 (17.3) | 15 (22.1) | 92 (23.7) | 115 (13.9) |  |
| 55-64 | 508 (16.7) | 325 (18.5) | 182 (14.2) | 4 (6.2) | 75 (19.3) | 103 (12.5) |  |
| 65+ | 629 (20.7) | 403 (22.9) | 226 (17.6) | 13 (20.4) | 78 (20.0) | 135 (16.3) |  |
| **Gender** |  |  |  |  |  |  |  |
| Male | 1443 (47.5) | 787 (44.7) | 657 (51.2) | 42 (64.1) | 184 (47.3) | 431 (52.0) | <0.0001 |
| Female | 1516 (50.0) | 946 (53.8) | 570 (44.4) | 23 (34.9) | 197 (50.7) | 350 (42.3) |  |
| Non-binary | 82 (2.7) | 26 (1.5) | 57 (4.4) | 1 (1.0) | 8 (2.1) | 48 (5.8) |  |
| **Race/Ethnicity** |  |  |  |  |  |  |  |
| Black NH | 350 (11.5) | 196 (11.2) | 154 (12.0) | 10 (14.8) | 38 (9.9) | 394 (47.6) | <0.0001 |
| White NH | 1794 (59.0) | 1122 (63.8) | 671 (52.3) | 23 (34.4) | 254 (65.3) | 106 (12.8) |  |
| Hispanic | 460 (15.1) | 176 (10.0) | 284 (22.1) | 21 (32.1) | 47 (12.1) | 216 (26.0) |  |
| Asian/Pacific Islander | 171 (5.6) | 82 (4.7) | 88 (6.9) | 4 (6.6) | 24 (6.2) | 60 (7.2) |  |
| Other | 268 (8.8) | 182 (10.3) | 86 (6.7) | 8 (12.2) | 25 (6.5) | 53 (6.4) |  |
| **Years of education** |  |  |  |  |  |  |  |
| Some HS and below | 353 (11.6) | 101 (5.7) | 252 (19.7) | 12 (18.6) | 23 (5.8) | 218 (26.2) | <0.0001 |
| HS Grad | 830 (27.3) | 501 (28.5) | 330 (25.7) | 27 (41.0) | 92 (23.7) | 211 (25.4) |  |
| Some college | 934 (30.7) | 594 (33.8) | 340 (26.5) | 11 (17.2) | 123 (31.5) | 206 (24.9) |  |
| College grad and above | 925 (30.4) | 563 (32.1) | 361 (28.2) | 15 (23.2) | 152 (39.0) | 195 (23.5) |  |
| **Household income** |  |  |  |  |  |  |  |
| Below 20K | 561 (18.4) | 253 (14.4) | 308 (24.0) | 18 (27.1) | 45 (11.5) | 245 (29.6) | <0.0001 |
| 20,000 - 60,000 | 941 (31.0) | 570 (32.4) | 371 (28.9) | 20 (29.7) | 107 (27.6) | 244 (29.5) |  |
| 60,000 - 100,000 | 568 (18.7) | 332 (18.9) | 236 (18.4) | 13 (19.3) | 101 (25.9) | 123 (14.8) |  |
| Above 100,000 | 460 (15.1) | 283 (16.1) | 177 (13.8) | 4 (5.5) | 80 (20.6) | 93 (11.2) |  |
| Prefer not to answer | 443 (15.6) | 274 (15.6) | 169 (13.2) | 8 (12.7) | 55 (14.3) | 105 (12.7) |  |
| DK | 69 (2.3) | 46 (2.6) | 23 (1.8) | 4 (5.8) | 1 (0.2) | 19 (2.3) |  |
| **Employed** |  |  |  |  |  |  |  |
| Yes | 1472 (48.4) | 798 (45.4) | 674 (52.5) | 32 (48.5) | 206 (52.9) | 437 (53.7) | <0.0001 |
| No/DK | 1570 (51.6) | 961 (54.6) | 610 (47.5) | 34 (51.5) | 183 (47.1) | 392 (47.3) |  |
| **Geographic region** |  |  |  |  |  |  |  |
| Northeast | 541 (17.8) | 271 (15.4) | 270 (21.0) | 11 (16.9) | 69 (17.7) | 190 (22.9) | <0.0001 |
| South | 1153 (37.9) | 709 (40.3) | 444 (34.6) | 28 (42.7) | 142 (36.4) | 275 (33.1) |  |
| Midwest | 630 (20.7) | 386 (22.0) | 243 (19.0) | 9 (13.8) | 80 (20.5) | 154 (18.6) |  |
| West | 718 (23.6) | 392 (22.3) | 326 (25.4) | 17 (26.6) | 99 (25.4) | 210 (25.3) |  |
| **Vaccination status** |  |  |  |  |  |  |  |
| Boosted | 1610 (52.9) | 809 (46.0) | 800 (62.3) | 24 (37.1) | 253 (65.1) | 522 (63.0) | <0.0001 |
| Fully vaccinated not boosted | 476 (15.6) | 329 (18.7) | 147 (11.4) | 15 (23.4) | 59 (15.3) | 72 (8.7) |  |
| Not vaccinated | 957 (31.5) | 620 (35.3) | 337 (26.3) | 26 (40.0) | 76 (19.6) | 235 (28.3) |  |
| **Prior COVID since March 2020** |  |  |  |  |  |  |  |
| Never | 1554 (51.1) | 1112 (63.2) | 442 (34.5) | 33 (49.7) | 199 (51.2) | 211 (25.4) | <0.0001 |
| Once | 808 (26.6) | 359 (20.4) | 451 (35.1) | 24 (37.0) | 117 (30.0) | 310 (37.4) |  |
| More than once | 343 (11.3) | 69 (3.9) | 274 (21.4) | 2 (2.4) | 24 (6.1) | 249 (30.0) |  |
| Never tested positive but think they had COVID | 335 (11.0) | 218 (12.4) | 116 (9.1) | 7 (10.9) | 50 (12.8) | 60 (7.2) |  |
| **Hybrid immunity** |  |  |  |  |  |  |  |
| Vaccine- and infection-induced | 987 (32.4) | 402 (22.9) | 585 (45.6) | 15 (22.4) | 145 (37.2) | 425 (51.3) | <0.0001 |
| Vaccine-induced only | 1099 (36.1) | 737 (41.9) | 362 (28.2) | 25 (38.1) | 168 (43.2) | 169 (20.4) |  |
| Infection-induced only | 501 (16.5) | 245 (13.9) | 256 (20.0) | 18 (27.9) | 45 (11.6) | 193 (23.3) |  |
| No immunity | 456 (15.0) | 375 (21.3) | 81 (6.3) | 8 (11.7) | 31.1 (8.0) | 42 (5.1) |  |
| **Comorbidities** |  |  |  |  |  |  |  |
| Yes | 1104 (36.3) | 584 (33.2) | 520 (40.5) | 20 (30.0) | 134 (34.6) | 366 (44.1) | <0.0001 |
| No | 1938 (63.7) | 1174 (66.8) | 764 (59.5) | 46 (70.0) | 255 (65.4) | 463 (55.9) |  |
| **Any vulnerability\*** |  |  |  |  |  |  |  |
| Yes | 2106 (69.2) | 1243 (70.7) | 863 (67.3) | 47 (71.4) | 232 (59.6) | 584 (70.5) | 0.04 |
| No | 936 (30.8) | 515 (29.3) | 420 (32.8) | 19 (28.6) | 157 (40.4) | 245 (29.5) |  |
| **Health insurance** |  |  |  |  |  |  |  |
| Yes | 2398 (78.8) | 1367 (77.8) | 1031 (80.3) | 53 (79.8) | 336 (86.3) | 643 (77.6) | 0.09 |
| No | 644 (21.2) | 391 (22.2) | 253 (19.7) | 13 (20.3) | 53 (13.7) | 186 (22.5) |  |

\*Aged 65 or older OR >1 comorbidity OR unvaccinated
\*\*Compares those who test at all with those who do not test at all

### Long COVID

Among respondents who had prior SARS-CoV-2 who also reported their most recent SARS-CoV-2 infection was more than 4 weeks prior to the survey (34.1% or 86,503,616 U.S. adults), an estimated 21.5% (95% CI 18.2, 24.7) reported long COVID symptoms (Table 3). Long COVID prevalence estimates varied across socio-demographic characteristics. Sex-standardized prevalence of long COVID was higher among respondents who were aged 35 - 44 (27.6%, 95% CI 19.3, 37.8), and age-standardized prevalence of long COVID was higher among female (27.4%, 95% CI 22.8, 32.6). Age- and sex-standardized prevalence of long COVID was higher among respondents who were Black non-Hispanic (27.3%, 95% CI 17.0, 41.0), unemployed (27.5%, 95% CI 20.6, 35.7) or had comorbidities (32.8%, 95% CI 25.9, 40.5). The standardized prevalence of long COVID was lower among respondents that were 65+ (14.8%, 95% CI 10.8, 19.9), male (15.5%, 95% CI 11.9, 20.2), or uninsured (14.0, 95% CI 7.7, 20.3).

**Table 3.** Prevalence and characteristics of U.S. adults with long COVID, July 2022.

|  | **Had COVID**<br>**more than**<br>**one month**<br>**ago** | **Long**<br>**COVID** | **Crude**<br>**Prevalence of**<br>**Long COVID** | **Age and sex**<br>**direct-**<br>**standardized**<br>**prevalence of**<br>**long COVID\*** | **Crude**<br>**prevalence**<br>**ratio (PR)** | **Adjusted**<br>**prevalence**<br>**ratio (aPR)\*\*** |
| --- | --- | --- | --- | --- | --- | --- |
|  | **N (%)** | **N (%)** | **% (95% CI)** | **% (95% CI)** | **PR (95% CI)** | **aPR (95% CI)** |
| **Total** | 1036 (100.0) | 222 (100.0) | 21.5 (18.2, 24.7) |  |  |  |
| **Age** |  |  |  |  |  |  |
| 18-24 | 70 (6.8) | 17 (7.8) | 24.8 (7.5, 42.0) | 18.6 (8.8, 35.2) | 1.1 (0.7, 1.7) | 1.2 (0.7, 1.9) |
| 25-34 | 250 (24.2) | 58 (26.1) | 23.2 (15.2, 31.2) | 22.3 (15.2, 31.5) | Ref | Ref |
| 35-44 | 178 (17.2) | 43 (19.1) | 23.9 (15.5, 32.3) | 27.6 (19.3, 37.8) | 1.0 (0.7, 1.5) | 1.2 (0.9, 1.8) |
| 45-54 | 171 (16.5) | 37 (16.5) | 21.6 (13.9, 29.3) | 22.3 (15.6, 30.9) | 0.9 (0.7, 1.3) | 1.1 (0.7, 1.5) |
| 55-64 | 185 (17.9) | 41 (18.2) | 21.9 (15.5, 28.3) | 23.1 (17.2, 30.3) | 0.9 (0.7, 1.4) | 1.1 (0.8, 1.5) |
| 65+ | 182 (17.6) | 27 (12.2) | 14.9 (10.5, 19.2) | 14.8 (10.8, 19.9) | 0.6 (0.4, 1.0) | 0.7 (0.5, 1.1) |
| **Gender** |  |  |  |  |  |  |
| Male | 465 (44.9) | 72 (32.4) | 15.5 (11.6, 19.4) | 15.5 (11.9, 20.2) | Ref | Ref |
| Female | 528 (51.0) | 144 (64.8) | 27.3 (22.2, 32.4) | 27.4 (22.8, 32.6) | 1.8 (1.4, 2.3) | 1.8 (1.4, 2.3) |
| Non-binary | 43 (4.2) | 6 (2.8) | 14.2 (0.0, 31.8) | 13.3 (4.8, 32.1) | 0.9 (0.4, 1.9) | 0.9 (0.4, 1.9) |
| **Race/Ethnicity** |  |  |  |  |  |  |
| Black NH | 75 (7.2) | 19 (8.4) | 24.9 (11.8, 38.1) | 27.3 (17.0, 41.0) | 1.2 (0.8, 1.8) | 1.1 (0.7, 1.7) |
| White NH | 690 (66.6) | 148 (66.8) | 21.5 (18.0, 25.1) | 22.4 (18.9, 26.5) | Ref | Ref |
| Hispanic | 129 (12.5) | 22 (9.7) | 16.8 (5.4, 28.1) | 16.8 (9.0, 29.2) | 0.8 (0.5, 1.2) | 0.8 (0.6, 1.2) |
| Asian/Pacific Islander | 41 (4.0) | 7 (3.2) | 17.6 (0.0, 37.5) | 11.0 (3.4, 30.6) | 0.82 (0.4, 1.6) | 0.8 (0.4, 1.7) |
| Other | 101 (9.8) | 26 (11.8) | 26.0 (13.1, 38.9) | 29.0 (19.7, 40.6) | 1.2 (0.8, 1.7) | 1.4 (0.9, 1.9) |
| **Years of education** |  |  |  |  |  |  |
| Some HS and below | 68 (6.5) | 16 (7.1) | 23.3 (4.8, 41.7) | 18.1 (7.4, 38.0) | 1.3 (0.8, 2.1) | 1.2 (0.7, 1.9) |
| HS Grad | 289 (27.9) | 69 (31.0) | 23.9 (16.7, 31.0) | 22.7 (15.8, 31.5) | 1.3 (1.0, 1.8) | 1.2 (0.9, 1.6) |
| Some college | 331 (31.9) | 75 (33.9) | 22.8 (17.1, 28.6) | 23.1 (18.0, 29.1) | 1.3 (1.0, 1.7) | 1.2 (0.9, 1.6) |
| College grad and above | 349 (33.7) | 62 (28.0) | 17.8 (13.8, 21.9) | 19.8 (15.7, 24.6) | Ref | Ref |
| **Household income** |  |  |  |  |  |  |
| Below 20K | 134 (12.9) | 34 (15.4) | 25.6 (14.8, 36.4) | 17.9 (11.5, 26.9) | 1.6 (1.0, 2.4) | 1.5 (0.9, 2.3) |
| 20,001 - 60,000 | 332 (32.0) | 83 (37.2) | 24.9 (18.6, 31.2) | 25.0 (19.1, 31.9) | 1.5 (1.1, 2.2) | 1.5 (1.0, 2.1) |
| 60,001 - 100,000 | 213 (20.6) | 48 (21.8) | 22.7 (15.9, 29.5) | 23.2 (17.1, 30.5) | 1.4 (0.9, 2.1) | 1.3 (0.9, 2.0) |
| Above 100,000 | 197 (19.0) | 32 (14.4) | 16.3 (10.2, 22.4) | 20.9 (14.5, 29.2) | Ref | Ref |
| Prefer not to answer | 160 (15.4) | 25 (11.1) | 15.5 (8.2, 22.7) | 15.6 (9.0, 25.8) | 1.0 (0.6, 1.5) | 1.0 (0.6, 1.7) |
| **Employed** |  |  |  |  |  |  |
| Yes | 548 (52.9) | 123 (55.3) | 22.4 (17.8, 27.1) | 24.3 (20.0, 29.3) | 1.1 (0.9, 1.4) | 1.1 (0.9, 1.4) |
| No/DK | 488 (47.1) | 99 (44.7) | 20.4 (15.8, 25.0) | 27.5 (20.6, 35.7) | Ref | Ref |
| **Geographic region** |  |  |  |  |  |  |
| Northeast | 187 (18.0) | 29 (13.2) | 15.8 (10.1, 21.4) | 17.2 (11.8, 24.3) | Ref | Ref |
| South | 402 (38.8) | 92 (41.5) | 23.0 (17.8, 28.2) | 22.6 (18.1, 27.9) | 1.5 (1.0, 2.1) | 1.5 (1.0, 2.1) |
| Midwest | 245 (23.6) | 52 (23.6) | 21.4 (14.9, 28.0) | 20.3 (14.7, 27.3) | 1.4 (0.9, 2.1) | 1.4 (1.0, 2.2) |
| West | 203 (19.6) | 48 (21.6) | 23.7 (14.7, 32.7) | 21.7 (14.9, 30.5) | 1.5 (1.0, 2.3) | 1.5 (1.0, 2.3) |
| **Vaccination status** |  |  |  |  |  |  |
| Boosted | 448 (43.3) | 86 (38.7) | 19.2 (14.8, 23.5) | 20.9 (16.4, 26.2) | Ref | Ref |
| Fully vaccinated not boosted | 196 (18.9) | 49 (22.2) | 25.1 (16.9, 33.4) | 24.9 (18.2, 33.0) | 1.3 (1.0, 1.8) | 1.2 (0.9, 1.6) |
| Not vaccinated | 392 (37.8) | 87 (39.2) | 22.2 (16.6, 27.9) | 22.8 (17.5, 29.1) | 1.2 (0.9, 1.5) | 1.1 (0.8, 1.4) |
| **Comorbidities** |  |  |  |  |  |  |
| Yes | 315 (30.4) | 96 (43.3) | 30.6 (24.5, 36.8) | 32.8 (25.9, 40.5) | 1.8 (1.4, 2.2) | 1.8 (1.4, 2.3) |
| No | 721 (69.6) | 126 (56.7) | 17.5 (13.7, 21.2) | 17.5 (14.2, 21.4) | Ref | Ref |
| **Health insurance** |  |  |  |  |  |  |
| Yes | 809 (78.1) | 190 (85.7) | 23.5 (19.8, 27.3) | 23.6 (20.1, 27.6) | 1.7 (1.2, 2.4) | 1.9 (1.3, 2.7) |
| No | 227 (21.9) | 32 (14.3) | 14.0 (7.7, 20.3) | 14.1 (0.9, 22.1) | Ref | Ref |

\*Direct standardized for the age and sex groupings based in the 2020 U.S. census, except for age (standardized for sex only) and gender (standardized for age only)
\*\*Models adjusted for gender and age

In age- and gender-adjusted models, we observed a higher prevalence of long COVID for females (versus male) (aPR: 1.8, 95% CI 1.4, 2.3), those with (versus without) comorbidities (aPR: 1.8, 95 % CI 1.4, 2.3), and those who were insured (versus not or unknown) (aPR: 1.9, 95% CI 1.3, 2.7). Compared to those who were currently boosted, we did not find statistically significant differences in prevalence of long COVID among those who were fully vaccinated but not boosted (aPR: 1.2, 95% CI 0.9, 1.6) and those who were not vaccinated (aPR: 1.1, 95% CI 0.8, 1.4).

## Discussion

We observed that 17.3% of adults in the U.S., or approximately 44.0 million adults, had SARS-CoV-2 infection during the two-week study period during the BA.5 surge in late June and early July 2022, much more than cases detected by case-based surveillance. This was at a time when the more transmissible BA.5 subvariant made up an estimated 57% of all cases in the week ending July 2, 2022.^2^ Importantly, there were major disparities in SARS-CoV-2 prevalence along the lines of social determinants of health, underscoring the long-standing inequities in SARS-CoV-2 burden in the US. Additionally, an estimated 18.6 million of the 86.5 million adults in the U.S. who reported having had SARS-CoV-2 at least once 4 or more weeks prior to the survey reported lingering symptoms consistent with long COVID. Disparities in SARS-CoV-2 infection during the 2-week period of our study portend a major contribution to subsequent disparities in long COVID burden. Our study provides important context to both the true burden of SARS-CoV-2 and its epidemiologic, sociodemographic, and geographic distribution.

The estimate of 44 million adults with SARS-CoV-2 infection during the two week study period, June 16 -July 2 is significantly higher than the 1.8 million cases in the official CDC case counts during the same period, which reported only ‘confirmed’ cases based on positive cases from nucleic acid amplification tests (NAAT)^13^. Our study underscores the extent to which reliance only on confirmed and reported cases contribute to the vast underestimation of the true burden of infection during surges. The degree of underestimation is likely increasing with time^4^. A few studies have examined the performance of case-based passive surveillance for SARS-CoV- 2^14,15^ but a broader evaluation of SARS-CoV-2 surveillance systems in relation to current public health goals^16^ is important.

Similar to prior findings using the same methodology^17,18^, we estimated a higher prevalence of SARS-CoV-2 infection among those who were vaccinated and boosted compared with those who were fully vaccinated but not boosted and those who were unvaccinated. Since vaccines and boosters provide limited protection against *infection* with omicron variants compared with prior strains^19^, these differences are likely due to differences in SARS-CoV-2 exposures and health behaviors between the two groups. Differences could also be partly explained by the higher testing rate among vaccinated and boosted persons, which when done for screening purposes, could result in greater detection of asymptomatic infection. These findings have important implications for observational (test negative) vaccine effectiveness (VE) studies, which are confounded by differences in exposure/behavior, testing behavior, and prior COVID between those vaccinated/boosted and unvaccinated.

We estimated 32.4% of adults had ‘hybrid immunity’ at the time of our survey. When we assessed the impact of both vaccination status and prior SARS-CoV-2 infection on risk of infection, we found that those with hybrid immunity had higher SARS-CoV-2 prevalence (29.2%), compared to those who had infection-induced immunity (17.0%), those who had vaccine-induced protection only (8.3%), and those who were SARS-CoV-2 naïve (5.8%). These findings suggest that prior infection, more so than vaccinations, may be an important marker for exposure risk during surges (e.g., workplace, household) and may also reflect a lower perceived risk for infection/reinfection. While there is evidence showing that prior infection may be negatively associated with decision to get the SARS-CoV-2 vaccine^20^, the possible role of past and more recent SARS-CoV-2 infections (including their timing) in reducing the adoption of personal risk mitigation measures during surges needs to be further examined.

Ascertaining the various demographic and socio-economic characteristics that underpin the current risk profile of SARS-CoV-2 exposure and infection remains important. As of February 2022, anti-nucleocapsid (anti-N) seroprevalence estimates suggest that more than half the U.S. population has had a prior SARS-CoV-2 infection^21^. Although SARS-CoV-2 primary vaccine and booster vaccination rates have stagnated^22^, based on a nationwide survey of blood donors, approximately 95% of the US population has some type of immunity (i.e., due to prior infection, vaccinations, or both)^13^. With the continued circulation of SARS-CoV-2 sub-variants across the country, recently acquired hybrid immunity is likely to play an increasingly important role in determining the potential impact of future surges on severe outcomes.

We also found that among those who had tested, a substantial proportion had tested exclusively with at-home rapid antigen tests (30%) which are not captured via case-based passive surveillance. Exclusive at-home testing was highest among non-Hispanic White adults, those with college and above levels of education, and those who have household income above $60,000. It was lowest among non-Hispanic Blacks and those with below high-school education. These findings are consistent with another study on the uptake of at-home test use^3^. We also found that 39.6% of our respondents have tested with at-home rapid tests, which is higher than a previous estimate of 20% of adults who tested with an at-home test during the BA.1 surge period^3^. The differential use of at-home tests by demographic characteristics and the increase in use of at-home testing over time provides insights about the groups in which infections are likely being undercounted by routine case-based surveillance.

Among respondents with SARS-CoV-2 infection more than four weeks ago, an estimated 21.5% reported currently having long COVID symptoms. Consistent with our estimates, the US Household Pulse Survey (HPS), an online survey sampling households which began collecting information on long COVID in June 2022, estimated 18.9% (95% CI 17.9, 19.8) of U.S. adults were currently experiencing long COVID.^9^ Our study, which was not restricted to persons accessing medical care, observed a lower prevalence of long COVID among the oldest (65+ years) versus younger age groups than reported elsewhere in the literature.^23,24^ This may be due to an overrepresentation of hospitalized or care-seeking patients in long COVID studies versus non-hospitalized populations. It could also reflect SARS-CoV-2 infection mitigation behaviors, such as mask wearing of social distancing, as well as higher early uptake of vaccines and boosters in older US residents, or survival bias (i.e., younger people are more likely to survive COVID-19 enabling them to report long symptoms of prior infection).

Our study has some limitations worth noting. First, our survey may have overestimated SARS-CoV-2 prevalence and provider testing if those with SARS-CoV-2 infection were more likely to participate in the survey. While potential survey participants were not aware of the survey content before deciding to participate, it may be that those who were positive were more likely to complete the survey. It is also possible that participants inadvertently recalled and reported positive tests that were beyond the 14-day study period (recall bias). In addition, some people test multiple times over a period of days or weeks with providers after their initial positive test^25^, and subsequently, many can expect positive PCR and antigen test results for 10 or more days^26,27^. This could have caused some people who were diagnosed prior to the study period to have positive tests during the study period which could have inflated our prevalence estimates relative to official case counts.

In addition, our case definition would likely capture a subset of the estimated 20-30% of individuals whose SARS-CoV-2 infection may remain asymptomatic throughout their infection^28^ as well as those who were symptomatic but were not aware of a close contact. To avoid confusion with acute COVID symptoms, we did not assess long COVID among respondents whose most recent SARS-CoV-2 infection was within the past month. However, some of these individuals may have long COVID from an earlier SARS-CoV-2 infection, which would result in an underestimation of the prevalence of long COVID. Finally, our survey did not include children or those whose primary language was not English or Spanish.

Strengths of our study include the representative and probability-based design of the survey, and the ability for the survey to reflect outcomes among those who do not access the healthcare system for SARS-CoV-2 and long COVID. Other strengths include the measurement of several important factors that are not currently available through routine surveillance, including outcomes among individuals vulnerable to COVID-19, hybrid immunity, and long COVID.

## Conclusions

We estimated a prevalence of SARS-CoV-2 during the BA.5 surge among adults in the U.S indicating substantial levels of SARS-CoV-2 circulation, much higher than official CDC case counts would suggest, with substantial disparities along the lines of social determinants of health. We also find that a significant proportion of U.S. adults with prior COVID (1 in five) report experiencing prolonged symptoms of long COVID. Our findings demonstrate the utility of population-representative surveys as an important surveillance tool to go alongside, and triangulate, with passive case reporting at an evolving stage of the U.S. pandemic.

## Data Availability

All data produced in the present study are available upon reasonable request to the authors.

## Acknowledgements

The authors wish to acknowledge the survey participants and Consensus Strategies for completing survey sampling and data collection.

## Appendix 1 (Survey design)

### Sampling Frame

A sampling frame of 254,297,978 Residents of the United States consisting of 105,469,157 mobile numbers with an additional 60,126,857 landlines. Two stratified proportionate randomized population-based samples were drawn for this study, n=90,000 mobile numbers and n=50,000 landlines. A National opt-in Online Panel provided by Consensus Strategies was used in the study. A total sample of n=3,042 was utilized with a +/- 3 percent margin of error. Data was collected June 30 - July 2, 2022.

### Multi-mode data collection design

Short message service (SMS) aka text messages were sent using SMS platform. The respondents were sent a personalized first name text message which included a link to the survey and an opt-out option. The respondents had the option to reply to the SMS text with any queries. Data was verified by IP address and scrubbed against the original survey sample.

Interactive voice response (IVR) aka robo-poll messages were sent to landlines. The respondents were able to answer the survey questions using the touch tone keypad on their phones.

The opt-in online panel was created by Consensus Strategies and participants were paid an incentive to complete the surveys of up to $2. Respondents were verified by payment information.

### Survey weighting

The survey was weighted using an iterative weighting method (raking) to marginal proportions of race, ethnicity, age, self-identified sex, and education by U.S. region. The samples (landline, online, mobile) were normalized at the region level based on sex, age, gender, education, race, and sample size then combined and weighted back based on the proportion of the region to the overall population and the other demographics. The sum of the weights equals the sample population (n=3,042). Demographic weights were created based on the American Community Survey 5-year estimates and <u>2020 US Census</u>. The inference population is 254,297,978 million adults in the U.S.

### Response rates

Our overall combined response rates across all modalities were 7.2%. The response rate was 6.2% for random digit dial to landline, 0.9% for cell phone, and 86.5% for opt-in online panel. The response rate reflects the proportion of complete respondents among eligible participants in the sampling frame. For context, we also included response rates for the Household Pulse Survey (HPS) in 2020 and 2022. While our response rates are comparable, the HPS methodology calculates the response rate based on completes and sufficient partial interviews, compared to our rates which are based on complete interviews (i.e., more conservative rates)

|  | *National COVID-19*<br>*Survey (June-July 2022)* |  |  |  | *HPS*<br>*2022*<br>*Jun*<br>*29-*<br>*Jul*<br>*11* | *HPS*<br>*2022*<br>*Jun*<br>*1-*<br>*Jun*<br>*13* | *HPS*<br>*2020*<br>*Week*<br>*1* | *HPS*<br>*2020*<br>*Week*<br>*2* | *HPS*<br>*2020*<br>*Week*<br>*3* |
| --- | --- | --- | --- | --- | --- | --- | --- | --- | --- |
| *Mode* | *Landline* | *Cell* | *Online*<br>*opt-in*<br>*panel* | *Combined* |  |  |  |  |  |
| *RR* | *6.2* | *0.9* | *86.5* | *7.2* | *5.7* | *6.2* | *3.8* | *1.3* | *2.3* |

## Appendix 2 (Survey questionnaire)

### Survey on recent COVID exposure, COVID infection, and testing behaviors in the United States

Hello, this is [interviewer] with a brief public policy survey. At no time will we try to sell you anything. We are just interested in your opinions, and you can drop out at any time.

To begin, what language would you like to take this survey in?

1. English
2. Español

#### The following questions will ask about COVID exposure in the past 2 weeks

1. In the past 2 weeks, have you experienced any COVID-like symptoms (e.g., 100 degrees fever or higher, chills, cough, sore throat, fatigue, headache, shortness of breath, congestion or runny nose, muscle aches, loss of smell or taste, nausea, or diarrhea)?
  a. Yes
  b. No
  c. Don’t know/not sure
2. In the past 2 weeks, were you aware of an exposure you had to someone who had COVID-like symptoms or tested positive for COVID-19?
  a. Yes
  b. No
  c. Don’t know/not sure

#### The following questions will ask about COVID testing in the past 2 weeks

3. In the past 2 weeks, have you taken an at-home rapid test for COVID-19? (a rapid at-home test allows you to collect your own sample and get results within minutes at home)
  a. Yes, Tested Positive
  b. Yes, Tested Negative
  c. No, I have not tested
4. In the past 2 weeks, have you taken a rapid antigen or PCR test for COVID-19 from a healthcare or testing provider?
  a. Yes, Tested Positive
  b. Yes, Tested Negative
  c. No, I have not tested [skip to 6]
5. *If tested with a healthcare provider*. When you tested with a healthcare or testing provider, which type of test did you receive?
  a. Rapid test/point of care test
  b. PCR test
  c. Both
  d. Not sure/don’t know
6. Prior to June 15th, 2022, did you ever have COVID-19 infection, either diagnosed by a healthcare or testing provider, or based on a positive at-home rapid test?
  a. Yes, once
  b. Yes, more than once
  c. No, but I am pretty sure that I had COVID
  d. No, I don’t think I have ever had COVID [skip to 8]
  e. Don’t know/not sure [skip to 8]
7. When was the last time you had COVID?
  a. Within the last month (if you currently have COVID, choose this option)
  b. 1-3 months ago
  c. 3-6 months ago
  d. 6-12 months ago
  e. >12 months ago
  f. Don’t know/not sure

#### Long COVID

8. Would you describe yourself as having ‘long COVID’, that is you experienced symptoms such as fatigue, difficulty concentrating, shortness of breath more than 4 weeks after you first had COVID-19 that are not explained by something else?
  a. Yes
  b. No [skip to 10]
  c. Don’t know/not sure [skip to 10]
9. Does this reduce your ability to carry-out day-to-day activities compared with the time before you had COVID-19?
  a. Yes, a lot
  b. Yes, a little
  c. Not at all
  d. Don’t know/not sure

#### Respondent Characteristics

10. Do you currently have any kind of health care coverage, including health insurance, prepaid plans such as HMOs, or government plans such as Medicaid or Medicare, or Indian Health Service?
  a. Yes
  b. No
  c. Don’t know/not sure
11. Do you have any of the following conditions that could increase the severity of COVID- 19: cancer, diabetes, obesity, COPD or lung disease, liver disease, heart disease, high blood pressure, a recent organ transplant, or an immunodeficiency)?
  a. Yes
  b. No
  c. Don’t know/not sure
12. Have you been fully vaccinated against COVID-19? [Either 2 doses of mRNA vaccine series (Moderna or Pfizer) or a single dose of Johnson and Johnson COVID-19 vaccine]
  a. Yes
  b. No (go to 14)
  c. Don’t know/not sure (go to 14)
13. If you have been fully vaccinated, have you also received at least one coronavirus booster?
  a. Yes, more than 5 months ago
  b. Yes, within the past 5 months
  c. No
14. If not fully vaccinated OR not boosted: Do you plan to get a vaccine dose or booster in the next two weeks?
  a. Yes
  b. No
  c. Don’t know/not sure
15. What is your age?
  a. 18-24
  b. 25-34
  c. 35-44
  d. 45-49
  e. 50-54
  f. 55-64
  g. 65-74
  h. 75 +
16. How do you currently identify your gender? Do you identify as …
  a. Male
  b. Female
  c. Gender non-binary
  d. other
17. Which one of the following would you use to describe yourself ?
  a. Latino/a, or of Hispanic or Spanish origin
  b. White
  c. Black or African American
  d. Asian, Native Hawaiian or Other Pacific Islander
  e. American Indian/Alaska Native
  f. More than one race
  g. Other
18. What is the highest grade or year of school you completed?
  a. Less than high school
  b. Grade 12 or GED (High school graduate)
  c. College 1 year to 3 years (Some college or technical school, associate degree)
  d. College 4 years or more (College graduate)
19. Are you currently employed for wages or salary?
  a. Yes
  b. No
  c. Don’t know/not sure
20. What is your household’s annual income?
  a. $20,000 or less
  b. Between $20,001 - $40,000
  c. Between $40,001 - $60,000
  d. Between $60,001 - $80,000
  e. Between $80,001 - $100,000
  f. Above $100,000
  g. Prefer not to answer
  h. Don’t know/not sure

